# Deep Chest: an artificial intelligence model for multi-disease diagnosis by chest x-rays

**DOI:** 10.1101/2024.12.05.24318531

**Authors:** Hakan Şat Bozcuk, Mehmet Artaç, Muzaffer Uğrakli, Necdet Poyraz

**Affiliations:** Medical Oncology, Private practice, Antalya, Turkey; Dept. of Medical Oncology, Necmettin Erbakan University, Konya, Turkey; Dept. of Radiology, Necmettin Erbakan University, Konya, Turkey

**Keywords:** Neural network model, computer vision systems, ai artificial intelligence, machine learning, chest, diagnosis, pulmonary nodule

## Abstract

**Background:** Artificial intelligence is increasingly being used for analyzing image data in medicine.

**Objectives:** We aimed to develop a computer vision artificial intelligence (AI) application using limited training material to aid in the multi-label, multi-disease diagnosis of chest X-rays.

**Methods:** We trained an EfficientNetB0 pre-trained model, leveraging transfer learning and deep learning techniques. Six thoracic disease categories were defined, and the model was initially trained on images sourced online and chest X-rays from a hospital database for training and internal validation. Subsequently, the model underwent external validation.

**Results:** In constructing and validating Deep Chest, we utilized 453 images, achieving an area under curve (AUC) of 0.98, sensitivity of 0.98, specificity of 0.80, and accuracy of 0.83. Notably, for diagnosing masses or nodules, the sensitivity, specificity, and accuracy were 0.97, 0.81, and 0.83, respectively. We deployed Deep Chest as a free experimental web application.

**Conclusions:** This tool demonstrated high accuracy in diagnosing both single and coexisting pulmonary pathologies, including pulmonary masses or nodules. Deep Chest thus represents a promising AI-based solution for enhancing diagnostic capabilities in thoracic radiology, with the potential to be utilized across various medical disciplines, especially in scenarios where expert support is limited.

## Introduction

Medical image classification has been one of the major triumphs in the field of AI. These image classification models have demonstrated success across various use cases, such as the utilization of magnetic resonance imaging (MRI) for identifying brain tumors, sorting images of blood leucocytes, diagnosing lung conditions through computed tomography (CT) scans, evaluating skin lesions, and performing differential diagnoses using histology images of breast cancer [1-7]. Pre-trained models have significantly streamlined the process of image classification by leveraging models already trained on general image classification tasks. Some prominent examples of these models include EfficientNet, Inception, ResNet, MobileNet, NasNet, and VGG, among others [8-13].

Transfer learning, a pivotal branch of AI, facilitates the application of knowledge gained from training a model in one specific domain to another domain, thereby reducing the time, effort, and resources needed [14]. Chest X-rays have been a staple in medical diagnostics for nearly a century and remain crucial for diagnosing common pulmonary pathologies [15]. However, accurately diagnosing or identifying multiple concurrent conditions using chest X-rays can be challenging and subjective, often requiring the expertise of seasoned radiologists who may not always be available.

In this study, we set out to develop a deep learning model capable of assisting clinicians in diagnosing single or multiple concurrent pulmonary conditions using chest X-rays, despite having limited training material. By focusing on the practical implementation of transfer learning and pre-trained models, we aimed to create a robust diagnostic tool that could operate effectively even in settings where expert radiological support is scarce. Our goal was to enhance the diagnostic process, making it more efficient and accessible, thereby improving patient outcomes not only in pulmonary healthcare but also in various other fields of medicine.

## Methods

### Selection of Chest X-rays for Training and Validation

To obtain the necessary chest X-ray images for our study, we utilized several sources. For training and internal validation, we sourced images from a randomly selected NIH chest X-ray dataset, various internet repositories, and real hospital chest X-rays from patients at the Oncology Department at Medical Park, Antalya. These patients were diagnosed with a range of health conditions [16]. We included chest X-rays from this oncology center only if the findings were confirmed by a thoracic CT scan within one week of the chest X-ray. To ensure data quality, any chest X-rays with faulty labeling were identified and excluded. The image file types allowed were JPG, JPEG, and PNG.

### Development of the Deep Chest Model (Training) and Internal Validation

We evaluated several pre-trained models, including EfficientNetB0 through EfficientNetB7, Inception V3, ResNet V2, and MobileNet V2, to compare their accuracy and computational resource requirements during the model fitting process. Approximately 10% of the chest X-ray images were reserved for internal validation. The model that exhibited the best performance with the least resource consumption was selected as the foundation for transfer learning.

Diagnostic categories were initially defined across 14 categories, but after observing that some categories did not yield accurate results, we consolidated them into 8, and ultimately 6 categories, which provided the highest internal validation accuracy. These final categories were “edema,” “mass or nodule,” “pneumonia,” “other pathologies,” “normal,” and “not a chest X-ray.”

To construct the AI model, we employed a sequential dense neural network structure from the Keras library for training and fitting the model. In addition to Keras, TensorFlow and NumPy libraries were utilized to develop the neural network structure, resulting in the Deep Chest model [17-19]. The entire project was implemented in Python, using the Google Colab Pro development environment. The model file was saved as an h5 file. During the later stages of development, hyperparameters were optimized to achieve the highest accuracy levels. The number of epochs used during training was determined based on validation AUC figures and the loss value obtained with each additional epoch.

### External Validation Stage

For external validation, an experienced radiologist from a teaching academic center (Poyraz N. from the Radiology Department of Necmettin Erbakan University, Konya) provided a set of 26 chest X-ray images from random cancer patients, along with their labels detailing thoracic diagnoses. Cancer patients were specifically chosen to include cases with likely concurrent pulmonary conditions. Additionally, we included a mini-set of 5 black and white and colored images of non-chest X-ray origin from the web to enrich the external cases, making the total number of images used in the external validation stage 31. Deep Chest was used to infer diagnoses from these images, and the results were compared with the labels provided by the radiologist or sourced from the web. We then calculated the accuracy, sensitivity, specificity, and AUC figures for both the internal and external validation cohorts.

### Development of Web Application

To enable chest X-ray uploading for inference, we developed a web application using the Streamlit library for Python [20]. The source code is hosted on a private repository on GitHub, and the web application is available online for free use [21]. This application allows users to upload chest X-rays and obtain diagnostic inferences from the Deep Chest model, thereby making our AI tool accessible for experimental use and further validation in clinical settings.

## Results

### Chest x-rays, pre-trained model, and hyperparameters

A total of 383 images for training, and 39 images for internal validation (9.2% of the total of 422) and 31 images for external validation are selected for building and validating the Deep Chest AI model. The total experience of inferencing with the Deep Chest model thus involved 453 images, and a total of 70 images were used to (internally and externally) validate the model. Among the pre-trained models tested, EfficientNetB1 to B7, Inception V3, ResNet V2, and MobileNet V2 yielded inferior accuracy scores or were more computing-resource-intensive compared to EfficientNetB0 (data not shown). So, EfficientNetB0 was selected for this study as the optimal pre-trained model, and the feature extractor function for EfficientNetB0 was set to “trainable”, to be able to adjust the model weights of the pretrained model with reference to the chest x-rays it was further trained with. For viewing the architecture of EfficientNetB0, see Figure 1.

**Figure 1.**
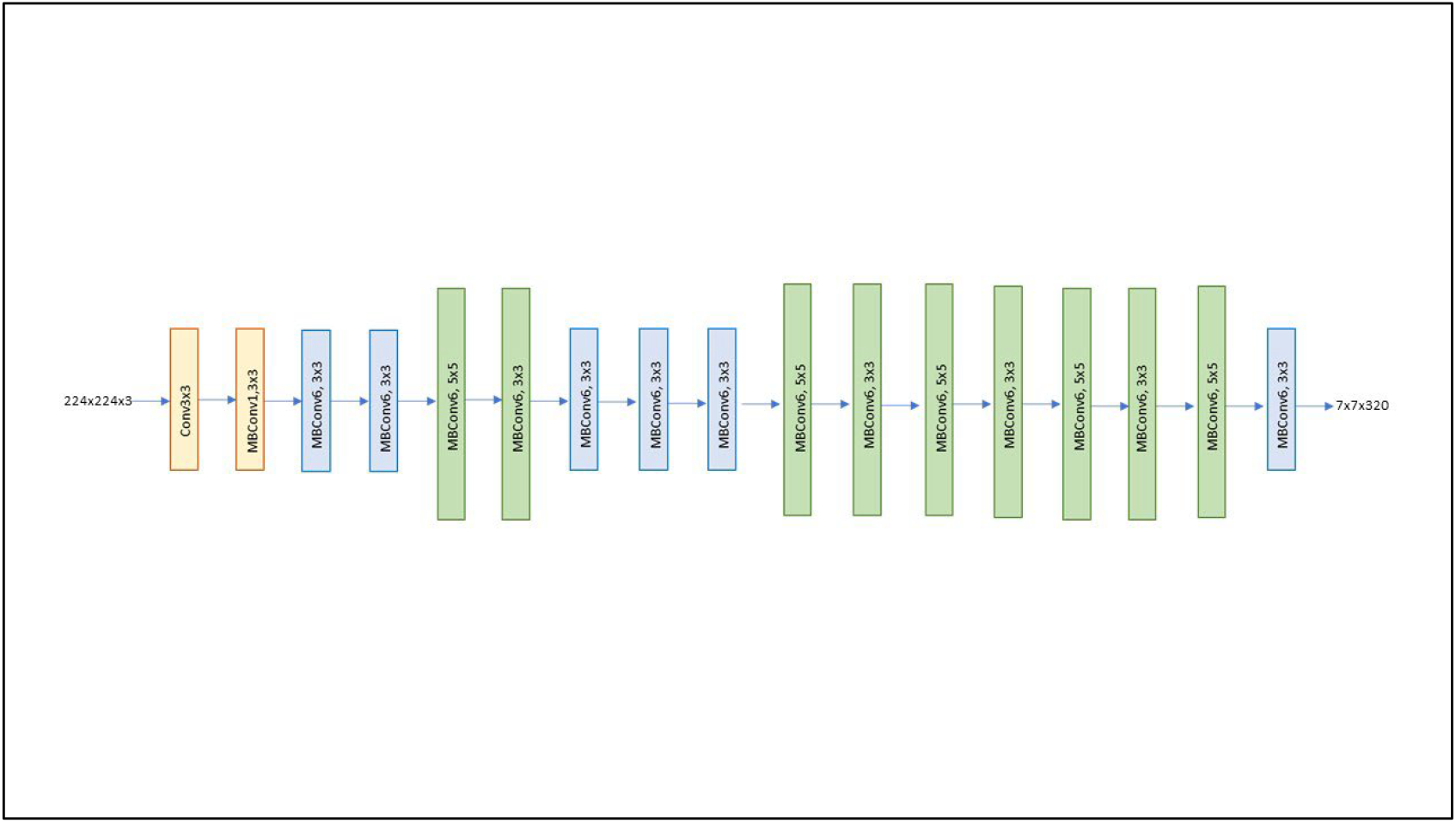
Architecture of EfficientNetB0.

A learning rate of 0.0001, epochs of 12, an activation function of sigmoid nature, batch size of 16, an optimizer function of Adam, and a loss type of “categorical_crossentropy” were selected as the optimal hyperparameters of the model.

### Model performance and clinical utility

During the training process, the Deep Chest model effectively learnt from the training material provided, as reflected in the significant decrease in both training and (internal) validation loss figures. The loss figure decreased by 74% in the training group, and by 47% in the validation group. See Figure 2 for the related plot. In parallel, the AUC figures increased from 0.66 to 0.98 in the training cohort, and from 0.70 to 0.92 in the validation cohort. Refer to Figure 3, to visualize how the model efficacy improved during training by viewing the change of AUC figures over epochs.

**Figure 2.**
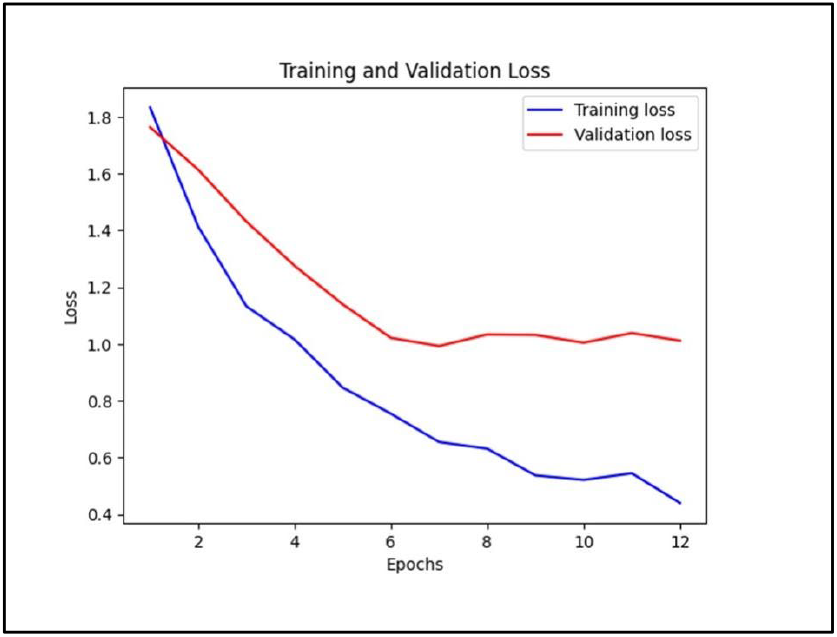
Loss during training and validation.

**Figure 3.**
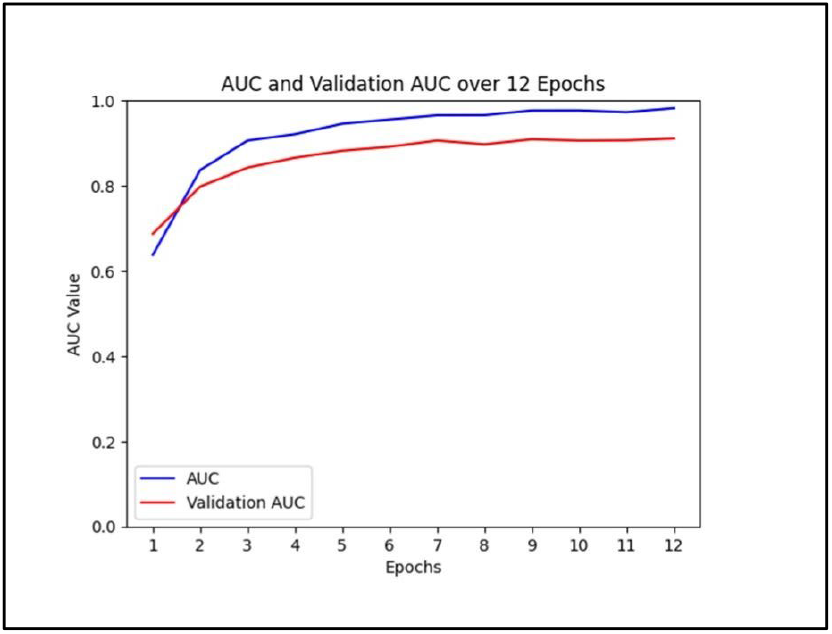
AUC during training and validation

At the training and internal validation stage, the overall model had an accuracy figure of 0.84. See Table 1 for a selection of efficacy metrics in different image cohorts; firstly, the training and internal validation combined, secondly, external validation, and thirdly, training, internal validation and external validation combined. With respect to the disease categories, accuracy figures ranged from 1.00 for the “not a chest x-ray” group, to 0.50 for the “pneumonia” group. Refer to Figure 4 for the breakdown of the efficacy metrics with respect to the categories in the training and internal validation combined cohort.

**Table 1.** Efficacy metrics in different cohorts.

|  | Training + internal<br>validation groups | External validation<br>group | Training + internal +<br>external validation<br>groups |
| --- | --- | --- | --- |
| Sensitivity | 1.00 | 0.92 | 0.98 |
| Specificity | 0.81 | 0.67 | 0.80 |
| Accuracy | 0.84 | 0.70 | 0.83 |
| AUC | 0.99 | 0.78 | 0.98 |

**Figure 4.**
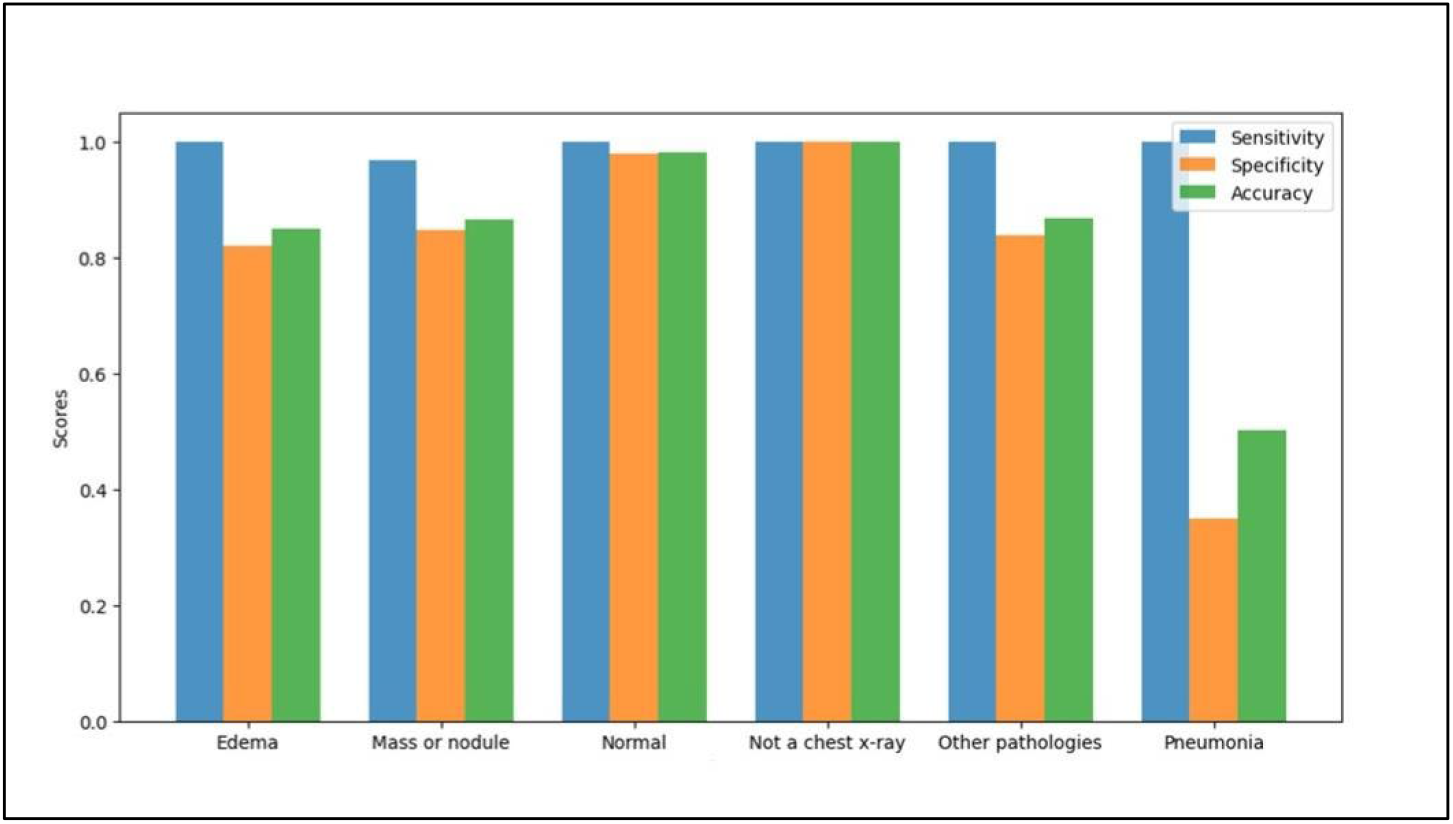
Efficacy metrics with respect to categories for the training and internal validation combined cohort.

For the external validation cohort, the accuracy measure was 0.70. When the categories were individually evaluated, again “not a chest x-ray” group had an accuracy of 1.00, whereas the lowest value was 0.42 for the “mass or nodule” group. Visit again Table 1 for the distribution of efficacy metrics in the external validation and other cohorts, and Figure 5 for the efficacy metrics in different categories for the external validation group.

**Figure 5.**
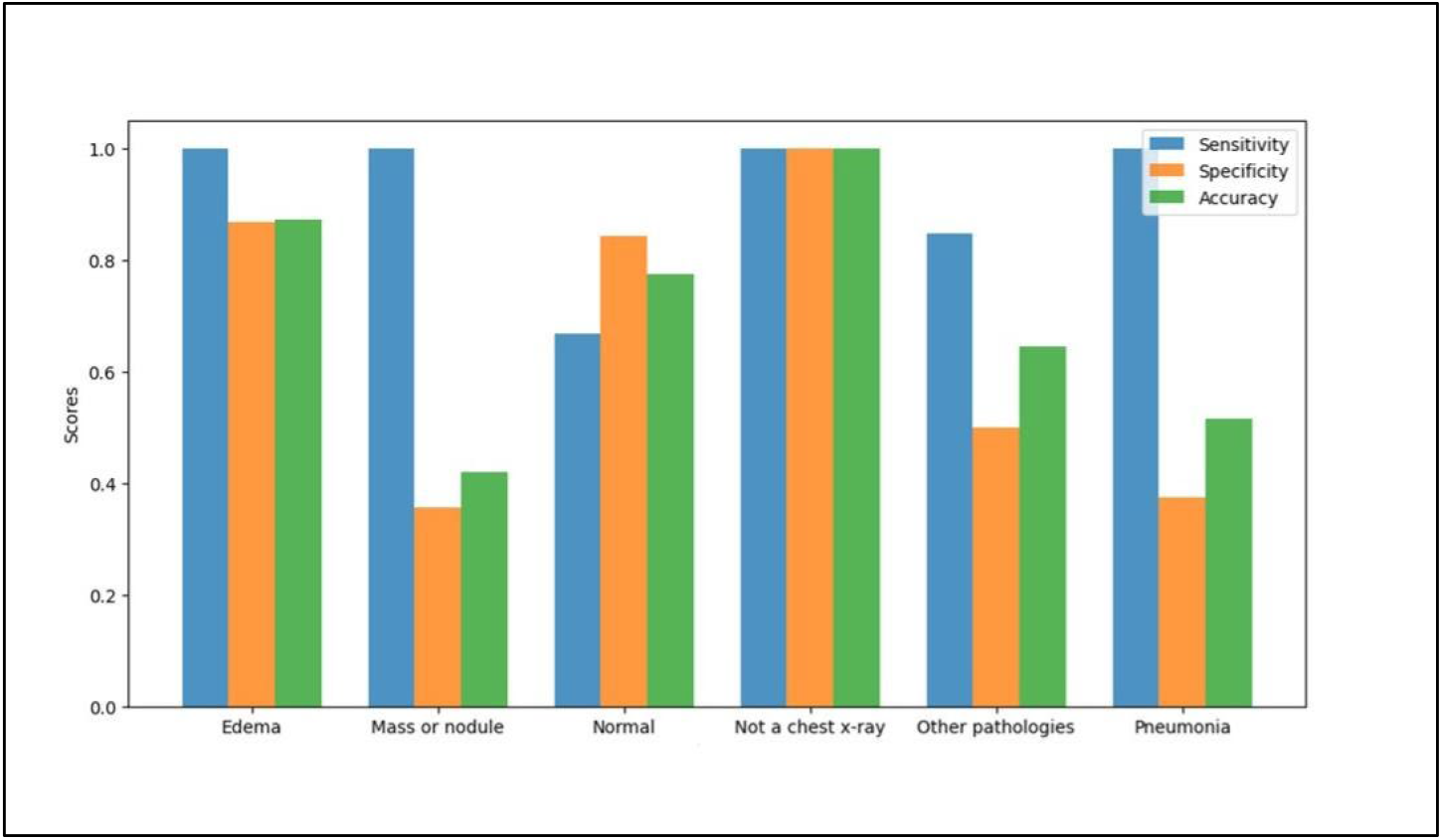
Efficacy metrics with respect to categories for the external validation cohort.

Lastly, when the whole Deep Chest experience is considered by combining the information from 3 cohorts (training, internal and external validation cohorts), resulting in a pool of 453 images, the accuracy value was 0.83. When the 6 categories were separately evaluated, the highest and the lowest accuracy figures were 1.00 and 0.50, again belonging to the “not a chest x-ray” and “pneumonia” categories. Go to Table 1 to see the efficacy details in the training, internal and external validation, and other cohorts. Figure 6 details efficacy details for this cohort of images.

**Figure 6.**
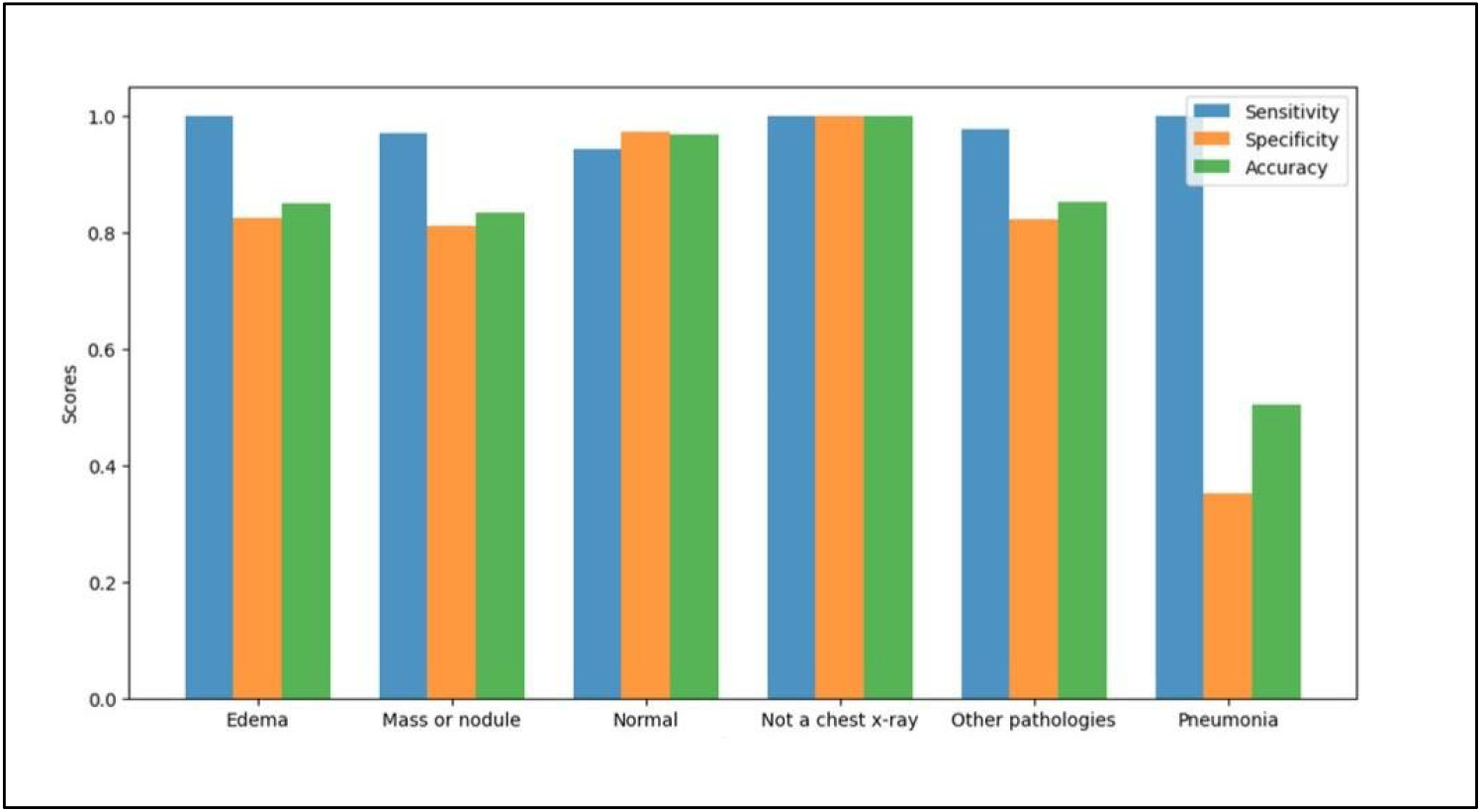
Efficacy metrics with respect to categories for the training, internal and external validation combined cohort.

When the AUC figures were considered, the overall model yielded 0.99, 0.78 and 0.98 in the training and internal validation combined, external validation, and training and internal validation and external validation combined cohorts. See Table 1 for the details of AUC figure, as well as other efficacy measures in various cohorts for the overall model. The distribution of AUC figures with respect to 6 disease categories is given in Figure 7 for the training and internal validation combined cohort, in Figure 8 for the external validation cohort, and in Figure 9 for the training, internal validation and external validation combined cohort, respectively.

**Figure 7.**
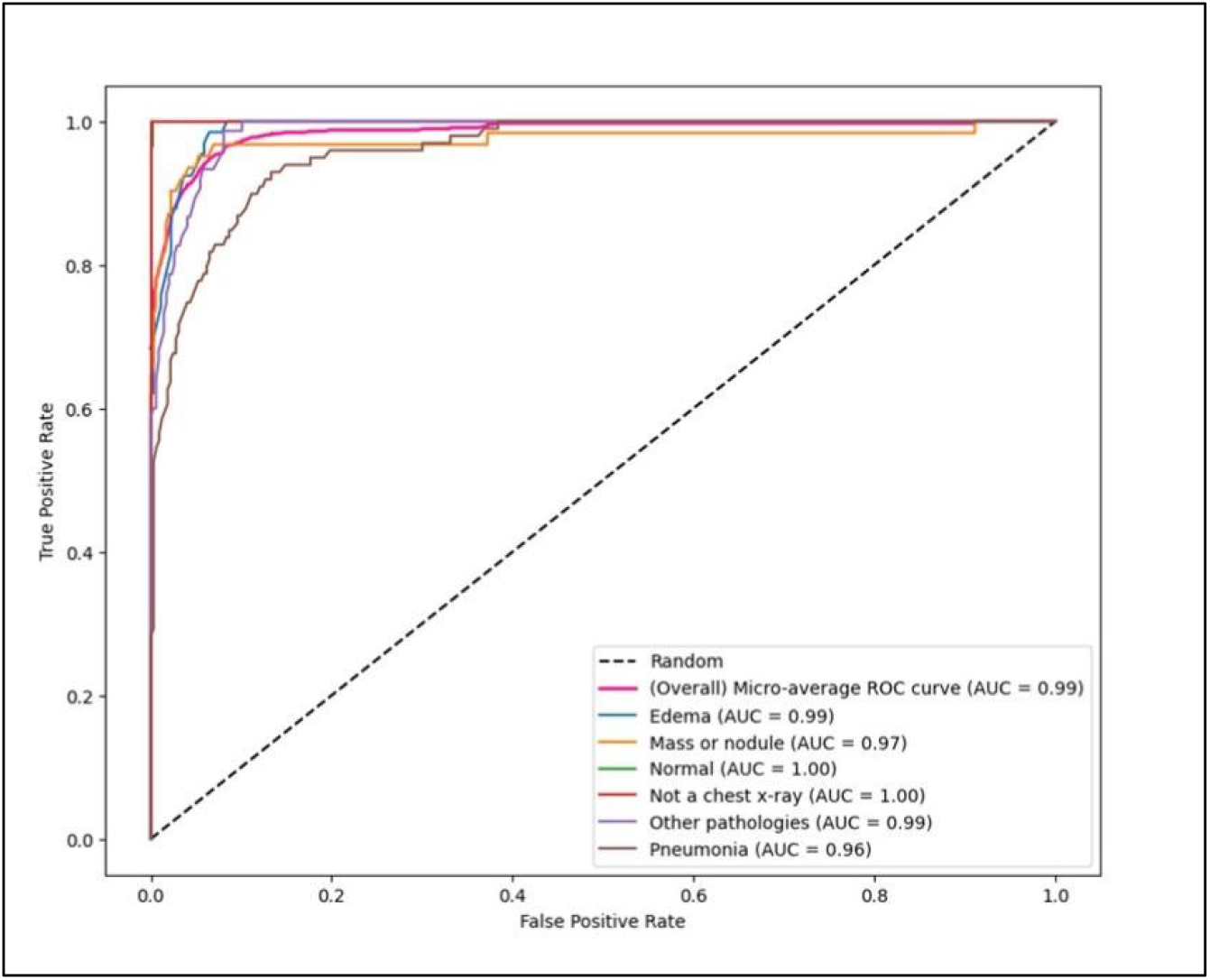
AUC figures with respect to categories for the training and internal validation combined cohort.

**Figure 8.**
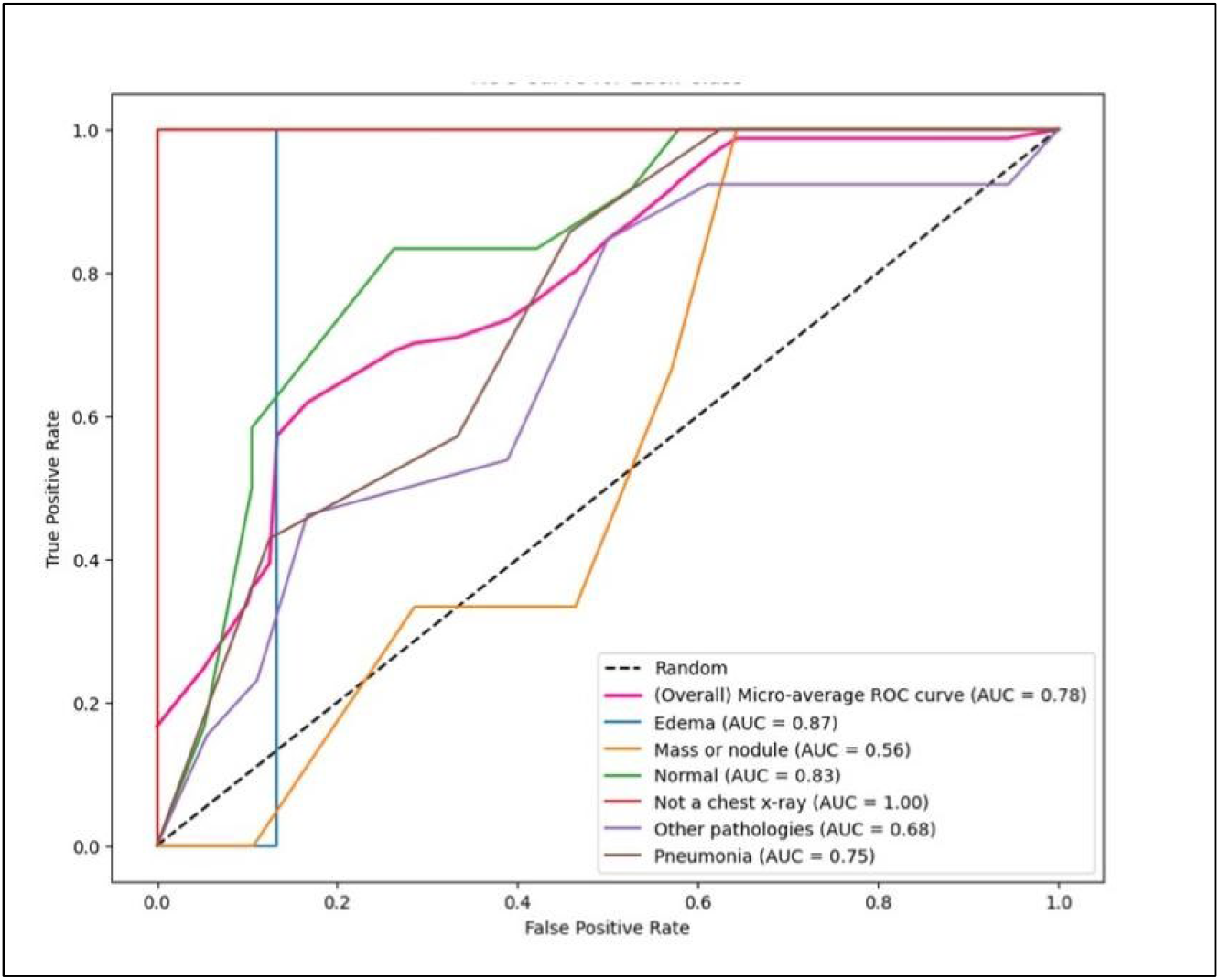
AUC figures with respect to categories for the external validation combined cohort.

**Figure 9.**
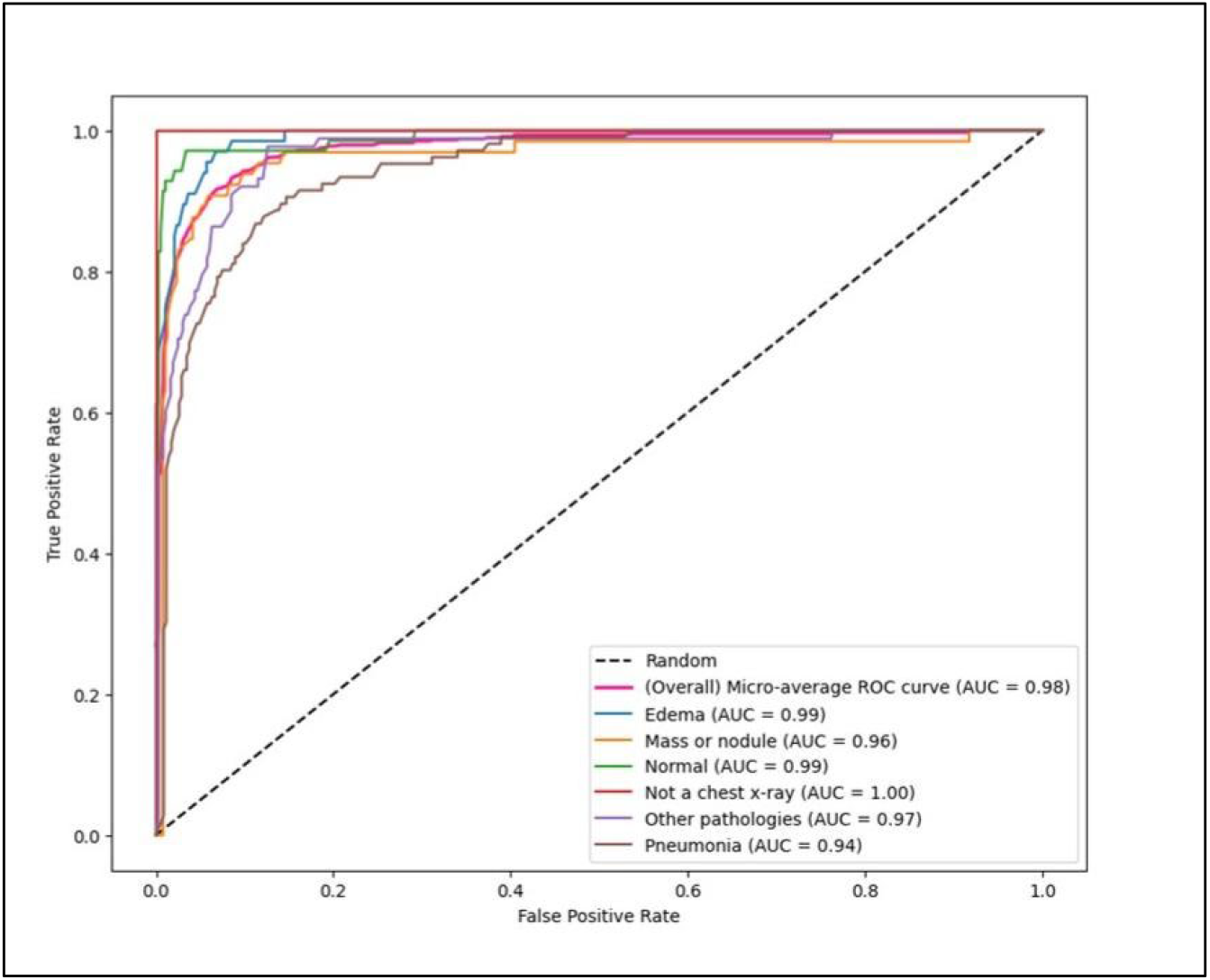
AUC figures with respect to categories for the training, internal validation and external validation combined cohort.

## Conclusions

Deep Chest achieved a good accuracy figure of 83% in the combination of 3 cohorts of images examined. This fact shows that Deep Chest has the potential to adequately predict the category of chest x-ray images. The model is perhaps more sensitive (98%) than it is specific (80%) as shown in the 3 cohorts combined. For the multilabel chest x-ray classification with deep learning, Pillai, et al. achieved an accuracy figure of 83% with the Inception model in 2022 [22]. Similarly, Rahmat, et al. used fast Region-based Convolutional Neural Network (R-CNN) architecture for chest x-ray image classification in 2019, and achieved an accuracy of 60% [23]. For the external validation cohort in our study, however, the accuracy by Deep Chest decreased to 70%. Therefore, Deep Chest appears to be more accurate than the model by Rahmat, et al, whereas, when compared to the model by Pillai, et al., Deep Chest looks to be as accurate, if our accuracy result in the 3 cohorts combined is considered, and less accurate, if only our external validation cohort with limited data is taken into account. In the literature, in addition to these multilabel models, chest x-ray classification has also been used for single label models, i.e., the case of distinguishing pulmonary nodules on chest x-rays, as shown by Nam, et al. in 2023 [24]. In their work, the identification of actionable pulmonary nodules has been increased by AI, with an odds ratio of 2.4. Thus, a number of AI models have already proven effective in chest x-ray diagnosis of multilabel or single label disease categories, and Deep Chest as shown with this work appears to be at least similarly effective as these models, while we employ a multilabel multiclass approach to chest x-ray classification, as we know the presence of coexisting diagnoses on chest x-rays is a common finding.

The types of AI methodology used for creating models for chest x-ray classification are various. For example, we and Pillai, et al. have used transfer learning, whereas Rahmat, et al. employed R-CNN [22,23]. Transfer learning, as we also used in our work, is a technique in machine learning, in which knowledge learned from a task is reused in order to boost performance on a related task. For the purpose of image classification, for example, knowledge gained while learning to recognize cars could be applied when trying to recognize trucks [25,26]. In our work, we used the knowledge gained while learning to recognize general objects in the ImageNet dataset, in the prediction of chest x-ray categories. To achieve that, we updated the weights in the pretrained EfficientNetB0 model, by using the information extracted from our collection of chest x-rays selected for training the AI model. Although our model is perhaps not convincingly more accurate than the currently available models, it is computationally “lighter”, since it requires much less training material to build the AI model. For Deep Chest, we only required 485 megabytes of training chest x-rays, as opposed to 11 gigabytes for the Pillai model, and 45 gigabytes for the Rahman model [22,23]. This is expected to lead to an improved training experience and a more efficient model building process. Some of the observed decrease in the magnitude of the training material required for building Deep Chest is partially attributable to the improved quality of the training material used, because, in addition to the images from the internet, we also employed carefully selected chest x-rays from our clinics; we used these chest x-rays in the model training process only if their findings were confirmed by the results of a chest CT scan within a week after an initial chest x-ray.

Looking at the diagnostic categories in our model, the sensitivity figures vary between 0.67 and 1.00 and the specificity figures are between 0.36 and 1.00. In detail, in the external validation cohort, “mass or nodule” category is associated with a sensitivity of 1.00 and a specifity figure of 0.36. In the training and validation cohort, the sensitivity and specifity measures are 0.97 and 0.85, respectively, indicating that the trained model is perhaps to some extent overfitted, and that in the external validation cohort, false positivity is prominent. Likewise, in the external validation cohort, “pneumonia” category is associated with the figures of specifity of 0.38 and sensitivity of 1.00, again indicating overdiagnosis of pneumonia cases during external validation. Therefore, Deep Chest cannot detect with enough specifity “mass or nodule” or “pneumonia” categories according to external validation, but when all cohorts are grouped together, Deep Chest exhibits remarkable efficacy in all categories, including ‘mass or nodule’ and ‘pneumonia,’ enabling the early diagnosis of pulmonary malignancies. However, other imaging modalities must be used to confirm or rule out malignancy due to Deep Chest’s high sensitivity but low specificity for that condition. These, taken together, imply that Deep Chest still has room for improvement at least for these 2 categories. This is potentially possible with providing further high-quality training material and experimenting with specific techniques to prevent model overfitting.

Additional work, using similar or more accurate methodologies or neural network architectures, making better use of transfer learning, trying different pre-trained models, employing more correctly labeled and confirmed chest x-rays for training or validation, is expected to improve the accuracy and specifity obtainable with Deep Chest. Artificial intelligence (AI) is transforming the field of medicine in unprecedented ways [27]. Within AI, computer vision stands out for its vast potential to enhance the diagnostic work-up of patients. Chest X-rays, a frequently used and cost-effective diagnostic modality, can now be analyzed with our Deep Chest model. This advancement allows for more accurate and timely diagnoses of pulmonary pathologies, including malignancies, across various medical disciplines. By leveraging transfer learning principles, we developed Deep Chest using a comparatively limited amount of training chest X-ray image data. This approach not only reduces the amount of data required but also simplifies and shortens the training process.

In short, we believe that AI models like Deep Chest will significantly aid clinicians in their routine work, providing valuable support in making diagnostic decisions. The model’s ability to efficiently handle and interpret medical images paves the way for its application in diverse clinical settings, especially where quick and accurate diagnoses are critical. However, continued efforts are necessary to refine these systems further. Future research should focus on developing more precise and reliable medical AI systems with complementary diagnostic capabilities. By integrating additional high-quality training data and exploring new methodologies, we can enhance the accuracy and robustness of AI-driven diagnostic tools, such as Deep Chest from this study, ensuring they meet the evolving needs of healthcare professionals and improve patient outcomes across the board.

## Data Availability

All data produced in the present study are available upon reasonable request to the authors

